# How suitable are clinical vignettes for the evaluation of symptom checker apps? A test theoretical perspective

**DOI:** 10.1101/2023.05.23.23290410

**Authors:** Marvin Kopka, Markus A. Feufel, Eta S. Berner, Malte L. Schmieding

## Abstract

**Objective:** To evaluate the ability of case vignettes to assess the performance of symptom checker applications and to suggest refinements of the methods of case vignette-based audit studies.

**Methods:** We re-analyzed the publicly available data of two prominent case vignette-based symptom checker audit studies by calculating common metrics of test theory. Furthermore, we developed a new metric, the Capability Comparison Score (CCS), which controls for different degrees of item difficulty of the set of cases each symptom checker evaluated. We then scrutinized whether applying test theory and the CCS alter the performance ranking of the investigated symptom checkers.

**Results:** In both studies most symptom checkers changed their rank order of triage capability when adjusting for item difficulty with the CCS. The previously reported triage accuracies commonly overestimated the capability of symptom checkers because they do not account for the fact that symptom checkers tend to selectively appraise easier cases. Many case vignettes in both studies showed insufficient (very low and even negative) values of item-total correlation.

**Conclusions:** A test theoretic perspective helps identify previously unreported limitations to the validity of case vignette-based symptom checker assessments and provides guidance on how to improve the quality of case vignettes and metrics for appraising their quality. A more elaborate metric, which accounts for item difficulty of vignettes an app did (not) evaluate, might prove more meaningful than accuracy alone for competitive assessment of symptom checkers. Our approach contributes to standardizing the methods to appraise symptom checker capability to yield more generalizable results.

## 1. Introduction

In recent years, symptom checkers were developed to aid laypersons in self-assessing their medical complaints ^1^. These tools, available as smartphone applications (“apps”) or web-browser based online applications, ask users a series of questions on their current medical complaints and list potential diagnoses (app-assisted self-diagnosis) and/or guidance on where to seek care as output (app-assisted self-triage) ^2^. As the use of such patient-facing clinical decision support systems has become increasingly popular ^3, 4^, it becomes an important task for researchers and regulators to develop methods to assess the accuracy and safety of their advice ^5^.

A recent systematic review revealed that currently the safety and accuracy of symptom checkers varies substantially between the individual apps ^6^, but also across the published studies assessing them. Among the possible reasons for this variation between studies are differences in the selected sample of apps (i.e., different apps or versions of them being assessed) and methodological differences, for example on how urgency levels were defined or on which types of complaints the apps were tested ^6^. For example, some authors evaluate symptom checkers on a broad variety of general symptoms ^1, 2, 7–11^, while others focus on symptoms or diseases from specific fields such as rheumatology or hepatitis C ^12–18^.

Another methodological difference is the source of input: either real-world patient histories ^8, 12, 13, 15, 17, 18^ or descriptions of fictitious patients ^1, 2, 7, 9–11, 16, 19^ serve as a basis for case vignettes with which symptom checkers are audited. Such case vignettes are usually created using various medical materials and are reviewed by multiple physicians to obtain a gold standard solution ^1^. While this approach originates from evaluating medical professionals ^20^ and transferring it to the evaluation of symptom checkers seems reasonable, the validity of this practice has been questioned ^21, 22^. For example, Haddad and Tylee ^23^ developed a test to evaluate school nurses’ recognition of depression and examined the clinical vignettes used for this purpose with a test theory approach. Though their findings indicate that most of the items used were appropriate for the authors’ purpose, some were unsuitable for the evaluation – because they were too easy to solve or did not correlate with the construct – although they were medically correct.

Because case vignettes are commonly used to competitively compare symptom checkers’ abilities and to audit their safety and accuracy, this paper aims to critically review this practice using a test theoretical perspective ^24^. Previous studies already noted limitations of case vignettes and recommended creating a benchmarking process to assess symptom checker accuracy ^25^ and to conduct studies “with greater methodological rigor and transparency” ^26^. By re-analyzing the data of previously published and highly influential studies, we take the first step by determining common metrics of test theory and using them to evaluate the ability of case vignettes to audit symptom checkers’ performance. The three questions we address are: a) using metrics from test theory, how suitable are case vignettes to assess symptom checker performance? b) if we account for low-quality vignettes, how does this change our interpretation of the currently published results on symptom checkers’ capabilities? c) in what way can a test theory perspective help advance the methodology for assessing symptom checkers’ capabilities? Our overall aim is to contribute to the standardization of evaluation methods which has been called for recently to improve the generalizability of studies on symptom checker performance ^27^.

As we consider triage advice from symptom checkers more important in regard to their safety and potential impact on healthcare delivery, we focus on triage recommendations from symptom checkers in this study, rather than their other use case of providing diagnostic suggestions.

## 2. Methods

### 2.1. Design

We conducted a secondary analysis of previously published data on triage accuracy of symptom checkers (test subjects) to calculate common test theoretical metrics of the case vignettes (test items) used in these studies.

### 2.2 Study Inclusion

This study aimed to include papers that are comparable to one another, are highly cited in this field and provide publicly available data. For this reason, only studies focusing on symptom checkers capable of handling a broad variety of symptoms and diseases were included, studies were further required to have used case vignettes and must have been cited at least 10 times by the time of our search (1^st^ Nov until 31^st^ Dec 2021).

Combinations of the terms ‘symptom checker’, ‘accuracy’, ‘reliability’, ‘self-triage’ and ‘self-diagnosis’ were entered into the database search engines Web of Science and Google Scholar, which led to the identification of 14 studies on symptom checker accuracy. Out of those, 8 did not focus on general symptoms, but specific symptoms or diseases such as COVID-19 ^12^ or orofacial pain ^19^. Of the remaining 6 studies, 1 did not use case vignettes ^8^ and 2 were cited no more than 10 times ^2, 9^. Finally, data of 1 study was not available upon request ^10^. Thus, 2 papers were included in the analysis for this study: One conducted by Hill and colleagues ^7^ and the other one conducted by Semigran and colleagues ^1^. Hill et al. examined 48 case vignettes with 16 unique symptom checkers providing triage advice (yielding a total of 688 case evaluations, as most symptom checkers were not able to assess all case vignettes) in 2020, while Semigran et al. tested 45 case vignettes with 15 symptom checkers providing triage advice (yielding a total of 532 case evaluations) in 2015. Eight of the evaluated symptom checkers were included in both studies. In its sample of 48 case vignettes, Hill et al. included modified versions of 30 case vignettes from the Semigran et al. study.

### 2.3. Metrics

Though numerous symptom checkers are available and included in the two studies, their number is rather low for a sample with which test items are validated and test-theory metrics are determined ^28^. Thus, we selected metrics from classical test theory, which can feasibly be calculated with a smaller number of data points. To satisfy this condition, we used two metrics from classical test theory: item difficulty and part-whole corrected item-total correlation ^24^. Based upon these metrics, we constructed new measures competitively re-evaluate the performance of the symptom checkers (Adjusted Accuracy, Capability Comparison Score).

#### 2.3.1. Metrics from Classical Test Theory on Test Items (Case Vignettes)

##### 2.3.1.1. Item difficulty

Item difficulty (ID) is a measure of how difficult a test item *i* is for the test subjects. In our case, this metric reflects the percentage of symptom checkers in a given study sample that were able to correctly solve the case vignette *i* and is calculated using the formula 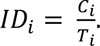 ID_i_ represents the item difficulty of case vignette *i*, C_i_ the absolute number of symptom checkers correctly solving case vignette *i*, and T_i_ the total number of symptom checkers providing advice to case vignette *i* (false or correct).

Thus, higher values indicate that the vignette was easier to solve (i.e., with a value of 1, all symptom checkers could solve the vignette), while lower values indicate a more difficult case vignette (i.e., with a value of 0.5, only half of the sample of symptom checkers evaluating this case vignette solved it correctly). This metric is relevant to create a balanced palette of test items, including both difficult and easy case vignettes to evaluate symptom checkers.

##### 2.3.1.2. Part-whole corrected item-total correlation

Item-total correlation (ITC) is a measure of item discrimination. For the purposes of our analysis this means that it reflects the degree to which correctly solving a specific case vignette (test item) is associated with a symptom checker’s accuracy on the remaining case vignettes. The part-whole corrected ITC is computed by calculating the accuracy for each symptom checker when the case vignette in question is omitted. Subsequently, the app’s suggestion on the omitted case vignette – coded as either true (correct suggestion) or false (incorrect suggestion) – is correlated across all symptom checkers with the respective accuracy excluding this case vignette. It can also be described as follows:

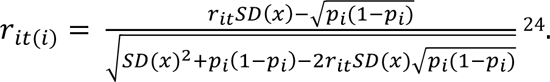

*r_it(i)_* represents the correlation of vignette *i* with the overall accuracy when vignette *i* is omitted, *SD(x)* the standard deviation of solving a vignette correctly, *p_i_* the probability that the vignette *i* was solved correctly, and *r_it_* the point biserial correlation of vignette *i* with the symptom checker’s accuracy *t* without omitting any vignette.

ITC values close to 0 indicate that solving vignette *i* correctly cannot predict the probability of a symptom checker to solve the remaining vignettes correctly. High values (up to a maximum score value of 1) indicate that vignette *i* is suitable for predicting the overall accuracy of symptom checkers. Negative ITC values indicate an inverse relationship between a symptom checker’s overall accuracy and accuracy regarding case vignette *i,* that is poor-performing symptom checkers tend to answer vignette *i* correctly, while high-performing symptom checkers tend to appraise case vignette incorrectly. Hence, ITC values close to 0 or negative indicate that the respective vignette is potentially inadequate to assess the performance of a symptom checker, having little informative value for a competitive comparison between symptom checkers. For the construction of test items in validated instruments, only items with ITC values above 0.2 are considered acceptable according to test theory, with values closer to 1 being preferable ^29^.

To further explore the construct validity of the measure triage accuracy we also determined each case vignette’s part-whole corrected item total correlation in a second manner: by measuring each vignettes correlation with symptom checkers’ accuracy on the remaining case vignettes of the same triage level (i.e., looking at the vignettes referring to emergency care, non-emergency care and self-care triage separately).

An ITC value cannot be calculated if a test item (here, case vignettes) is solved correctly by either all or none of the test subjects (here, symptom checkers). Such test items are also excluded from validated instruments as they provide no value for the competitive comparison between test subjects.

##### 2.3.1.3. Metrics Re-Evaluating the Test Subjects (Symptom Checkers)

To study whether the calculated values for item difficulty and part-whole corrected item-total correlation justify a re-interpretation of the results of the original studies, we created two new metrics. Both metrics revisit the competitive comparison of the symptom checkers’ triage capabilities. The first, adjusted accuracy, investigates how the results would change if shortcomings relating to part-whole corrected item total correlation in the vignette sample were considered. The second, the capability comparison score, explores how taking item difficulty into account changes the assessment of a symptom checker’s capability.

##### 2.3.1.4. Adjusted Accuracy

We used the part-whole corrected item-total correlation as foundation for a critical reflection of the informative value of the raw accuracy metric commonly used to appraise a symptom checker’s ability. In the two analyzed studies, accuracy represents the proportion of case vignettes that were solved correctly from cases the given symptom checker evaluated ^1, 7^. As most symptom checkers cannot evaluate all vignettes and differ with respect to the kinds of vignettes they can solve, their reported raw accuracy might be based on samples of case vignettes with different informative value (as measured by the case vignettes’ ITC values). Based on the ITC value of a vignette, we defined a new metric complementing this (raw) accuracy metric: adjusted accuracy. To calculate the adjusted accuracy, we included only vignettes with an ITC greater than 0.2, following the common practice in test theory to exclude items with negative and too low ICTs because they add noise rather than help to discriminate between high and low performing symptom checkers ^24^.

Thus, adjusted accuracy equals a symptom checker’s raw accuracy eliminating those case vignettes with low or questionable informative value. Taken together, these two measures of accuracy (raw and adjusted) can be interpreted as follows: when a symptom checker’s raw accuracy diverges from its adjusted accuracy, then its raw accuracy might be biased by unsuitable case vignettes leading to either an overestimation or underestimation of the symptom checker’s capability. This information might also be used to exclude low quality vignettes in future studies.

##### 2.3.1.5. Capability Comparison Score

Unlike in most tests to which test theory pertains, in symptom checker studies it is common that each app assesses a (slightly) different subset of test items (case vignettes), as most symptom checkers are not able to provide advice regarding all chief complaints featured in a given study’s pool of case vignettes: In the Semigran et al. study only 2/15 apps considered all case vignettes, 4/16 in the Hill et al. study respectively.

Item difficulty is therefore also insightful when comparing the raw accuracy values of symptom checkers that have evaluated different sets of case vignettes. For example, two symptom checkers might have similar raw accuracies, but one having evaluated and solved correctly less difficult cases than the other, arguably making it less capable than the other. To look into this, we determined for every symptom checker the mean (M) and standard deviation (SD) of item difficulty of the vignettes, which it a) evaluated and b) solved correctly, a subset of the former.

Based on these data we defined and calculated a Capability Comparison Score for each symptom checker. We defined the Capability Comparison Score (CCS) to enable direct comparisons between symptom checker performance by weighing the performance measure with the ID of the case vignettes a particular symptom checker solved. To obtain the CCS for a symptom checker, the IDs of correctly solved cases are summed up and the sum of the IDs of incorrectly solved cases are subtracted to penalize the symptom checker for incorrect answers. Lastly, it normalizes these values to limit the range of values to 0% and 100%. This can be described using the following formula:

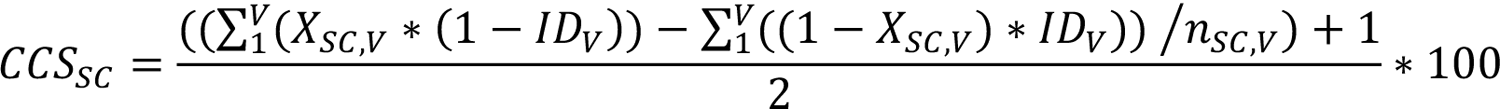

where CCS represents the Capability Comparison Score, SC the symptom checker, V the case vignette, X the test score (i.e., Boolean whether it was solved correctly), ID the item difficulty and n_SC,V_ the number of entered case vignettes in a symptom checker. A visual explanation of the formula components can be found in Figure 1.

**Figure 1.**
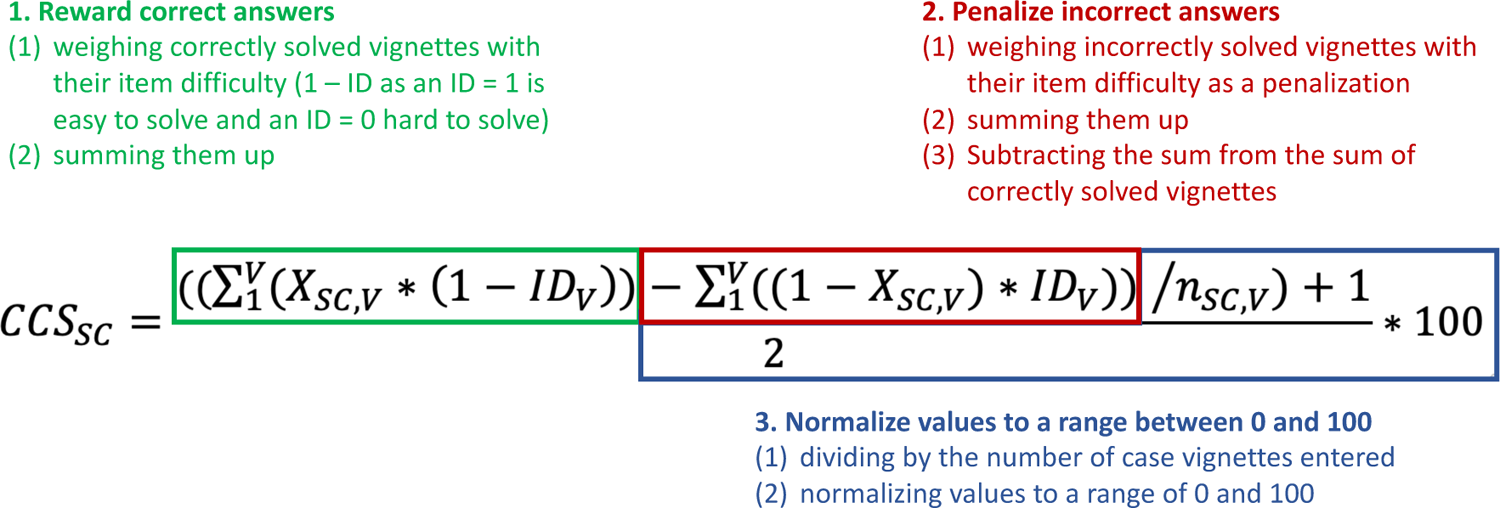
Explanation of the CCS formula components

For example, a symptom checker providing equally often correct and incorrect advice on case vignettes all with an ID of 0.5 would receive a CCS value of 50%, the same as the raw accuracy score of 50%. In comparison, a different symptom checker with the same raw accuracy of 50% but having assessed more difficult case vignettes all with IDs of 0.3 would achieve a higher CCS value (namely, 64). A third symptom checker with an accuracy of 50% having assessed case vignettes of lesser difficulty with IDs of 0.7 would achieve a lower CCS value (namely, 17).

### 2.4. Data Analysis

Data were analyzed and visualized using *R* version 3.6.1 ^30^ and the *tidyverse* packages ^31^. For descriptive analyses, the median (Mdn) and interquartile range (IQR) were calculated when data were not normally distributed. When data were normally distributed, we calculated mean (M) and standard deviations (SD). Distributions were visualized using raincloud plots ^32^.

Finally, we analyzed a subset of the data by including only vignettes used in both studies (*n* = 30) to examine the stability of the test theoretic metrics. A Pearson correlation of the metrics between the studies was calculated, the data points were plotted, and their relationship visualized using linear regression.

## 3. Results

### 3.1. Item Difficulty

While the case vignettes were more difficult in the Hill study than in the Semigran study, the spread of item difficulty was large and of about equal magnitude in both studies: For vignettes used by Hill et al., the item difficulty was *Mdn* = 0.455 (*IQR* = 0.412) and for those used by Semigran et al. *Mdn* = 0.538 (*IQR* = 0.452). Both studies include vignettes that either all or no symptom checker solved correctly (3/48 in the vignette sample by Hill et al., and 4/45 in the vignette sample from Semigran et al.). The distribution of item difficulties of the vignettes is visualized in Figure 2.

**Figure 2.**
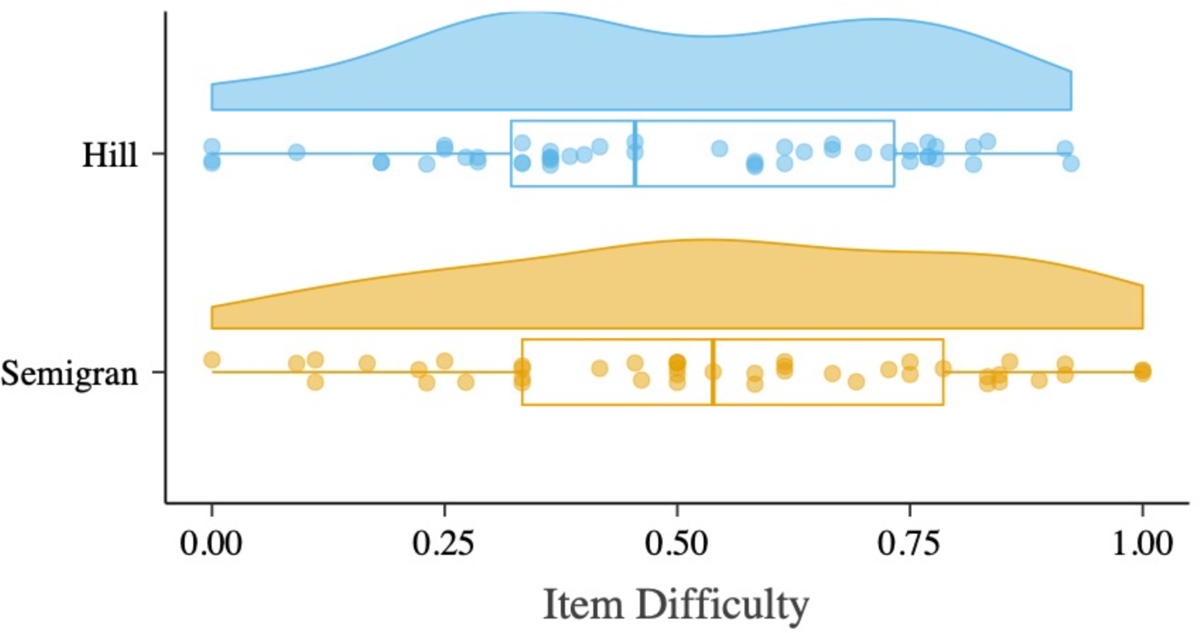
Distribution of Item Difficulty of Case Vignettes Tested Differentiating Between Included Studies

The symptom checkers in the Hill et al study all considered samples of case vignettes with a similar mean item difficulty: the mean ID for the app considering the most difficult sample of case vignettes, *HealthyChildren,* was 0.470, close to the mean ID for *EverydayHealth* and *Symptomate* (both 0.502), the apps considering the least difficult sample of case vignettes in the Hill study. This gap is more than 7 times greater in the Semigran et al study (minimum mean ID (*HealthyChildren*): 0.460, maximum mean ID (*Symptomate*): 0.692), see Table 1 and 2.

**Table 1.** Item Difficulty by Symptom Checker, Hill et al. Study

| App | Raw Accuracy | Item Difficulty of Entered Cases, M (SD) | Capability Comparison Score | Rank Based on Raw Accuracy | Rank Based on Capability Comparison Score |
| --- | --- | --- | --- | --- | --- |
| Healthdirect | 60.9% (28/46) | 0.495 (0.259) | 55.1% | 1 | 1 |
| UofM Health | 57.8% (26/45) | 0.494 (0.254) | 54.2% | 2 | 2 |
| Healthlink BC | 56.2% (27/48) | 0.491 (0.255) | 53.6% | 3 | 4 |
| Healthwise | 56.2% (27/48) | 0.491 (0.255) | 53.6% | 3 | 4 |
| Drugs.com Google Play | 56.1% (23/41) | 0.485 (0.252) | 53.8% | 5 | 3 |
| Drugs.com | 53.7% (22/41) | 0.485 (0.252) | 52.6% | 6 | 6 |
| Everyday Health | 51.6% (16/31) | 0.502 (0.244) | 50.7% | 7 | 7 |
| Symptomate | 48.4% (15/31) | 0.502 (0.244) | 49.1% | 8 | 13 |
| Isabel | 47.9% (23/48) | 0.491 (0.255) | 49.4 | 9 | 12 |
| Healthy Children | 47.1% (8/17) | 0.470 (0.280) | 50.0% | 10 | 8 |
| Hopkins all Children | 47.1% (8/17) | 0.470 (0.280) | 50.0% | 10 | 8 |
| St. Lukes Online | 47.1% (8/17) | 0.470 (0.280) | 50.0% | 10 | 8 |
| CHW | 47.1% (8/17) | 0.470 (0.280) | 50.0% | 10 | 8 |
| Symcat | 37.0% (17/46) | 0.497 (0.248) | 43.6% | 14 | 14 |
| Family Doctor | 29.8% (14/47) | 0.501 (0.247) | 39.8% | 15 | 15 |
| Doctor Diagnose | 25.0% (5/20) | 0.501 (0.246) | 37.4% | 16 | 16 |
| Total | 48.0% | 0.491 (0.252) | 49.6% |  |  |

*Note.* Since the number of entered cases varied for each symptom checker, Total represents the unweighted mean of all symptom checkers and the mean capability comparison score of values of all symptom checkers

### 3.2. Capability Comparison Score

The mean CCS for the symptom checkers assessed in the Hill study (49.6%, SD = 5.15%) is similar to their mean raw accuracy (48.0%, SD = 10.0%); in comparison, the mean CCS in the Semigran study (50.4%, SD = 6.85%) is lower than the mean raw accuracy (58.0%, SD = 12.8%), see Table 1 and Table 2. In line with this, all but three apps received a lower CCS than their raw accuracy in the Semigran study. In contrast, half of symptom checkers (9/16) achieved a higher CCS score than their raw accuracy in the Hill study, with 7 of 16 receiving a lower CCS.

**Table 2.** Item Difficulty by Symptom Checker Semigran

| App | Accuracy | Item Difficulty of Entered Cases, M (SD) | Capability Comparison Score | Rank Based on Raw Accuracy | Rank Based on Capability Comparison Score |
| --- | --- | --- | --- | --- | --- |
| HMS Family Health Guide | 80.0% (32/40) | 0.588 (0.265) | 60.6% | 1 | 2 |
| Healthy Children | 73.3% (11/15) | 0.460 (0.325) | 63.7% | 2 | 1 |
| Steps2Care | 71.4% (30/32) | 0.551 (0.278) | 58.2% | 3 | 3 |
| Symptify | 70.0% (28/40) | 0.558 (0.265) | 57.1% | 4 | 4 |
| Symptomate | 64.3% (9/14) | 0.692 (0.251) | 47.5% | 5 | 11 |
| Doctor Diagnose | 62.5% (10/16) | 0.681 (0.286) | 47.2% | 6 | 12 |
| Drugs.com | 59.5% (25/42) | 0.578 (0.265) | 50.9% | 7 | 6 |
| FreeMD | 59.1% (26/44) | 0.565 (0.268) | 51.3% | 8 | 5 |
| Family Doctor | 53.7% (22/41) | 0.537 (0.272) | 50.0% | 9 | 7 |
| Early Doc | 52.9% (9/17) | 0.573 (0.203) | 47.8% | 10 | 8 |
| NHS | 52.3% (23/44) | 0.566 (0.267) | 47.8% | 11 | 8 |
| Isabel | 51.1% (23/45) | 0.555 (0.274) | 47.8% | 12 | 8 |
| Symcat | 44.4% (20/45) | 0.555 (0.274) | 44.5% | 13 | 13 |
| Healthwise | 43.2% (19/44) | 0.557 (0.276) | 43.7% | 14 | 14 |
| iTriage | 32.6% (14/43) | 0.564 (0.275) | 38.1% | 15 | 15 |
| Total | 58.0% | 0.566 (0.270) | 50.4% |  |  |

*Note.* Since the number of entered cases varied for each symptom checker, Total represents the unweighted mean accuracy of all symptom checkers and the mean capability comparison score of values of all symptom checkers

When ranked by the CCS rather than the raw accuracy, 9 of 16 and 10 of 15 symptom checkers change their rank position in the Hill and the Semigran study, respectively. Most of these rank order changes occur in the mid-tier of apps, that is the two best or worst performing apps in each study are not affected much in terms of rank order changes.

### 3.3. Item-Total-Correlation

Distribution of Item-Total-Correlation Values

For some case vignettes, no item-total correlation could be calculated because either all symptom checkers solved them correctly or none did (Hill_AllCorrect_ = 0, Hill_NoneCorrect_ = 3, Semigran_AllCorrect_ = 3, Semigran_NoneCorrect_ = 1). The distribution of ITC values of the remaining vignettes is displayed in Figure 2 for both studies.

In the study conducted by Hill et al., 40% (18/45) of case vignettes for which an ITC could be calculated had a negative item-total-correlation. Those with a positive item-total-correlation had a Median of 0.431 (IQR = 0.368). Of these, 21 reached an item-total-correlation above 0.2 (*Mdn* = 0.517, IQR = 0.271).

In the study conducted by Semigran et al., 29.3% (12/41) of case vignettes for which an ITC could be calculated had a negative item-total-correlation. Those with a positive item-total-correlation had a Median of 0.425 (IQR = 0.236). Of these, 23 reached an item-total-correlation greater than 0.2 (*Mdn* = 0.450, IQR = 0.171. The distributions of item-total-correlations are visualized in Figure 3.

**Figure 3.**
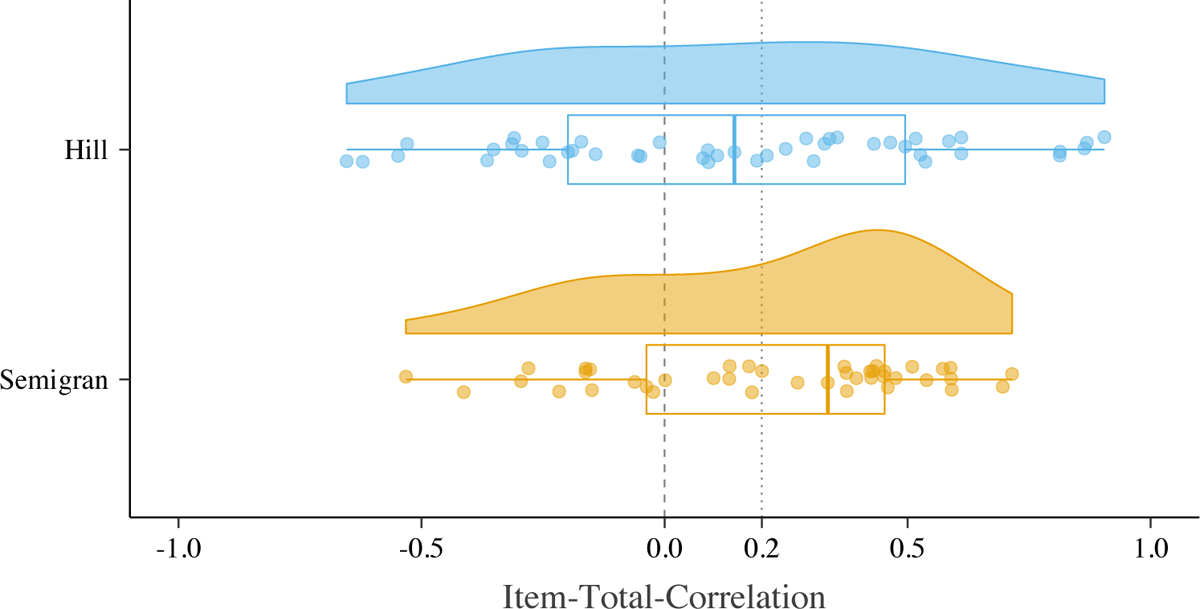
Density of Item-Total-Correlation by Study

Notably, all emergency case vignettes (15/15) in the Semigran study presented an ITC value below 0.2. In the Hill study, the majority of emergency case vignettes (7/13) had an ITC value greater than 0.2.

To aid the reader in the interpretation of ITC values and to doublecheck the logic of the ITC, Table 3 presents the mean raw accuracies of symptom checkers solving the case vignettes with the most extreme ITC values (highest positive, highest negative and lowest ITC) correctly and incorrectly: Concordant with ITC’s logic, the average raw accuracy of symptom checkers providing correct triage advice is lower than those solving it incorrectly for the vignette with the lowest ITC value, i.e., the otherwise highly performing apps fail on this case vignette, while the otherwise poorly performing apps do not. Conversely, the vignette with the highest ITC value was solved correctly by highly performing apps and incorrectly by apps of lower triage accuracy.

**Table 3.**
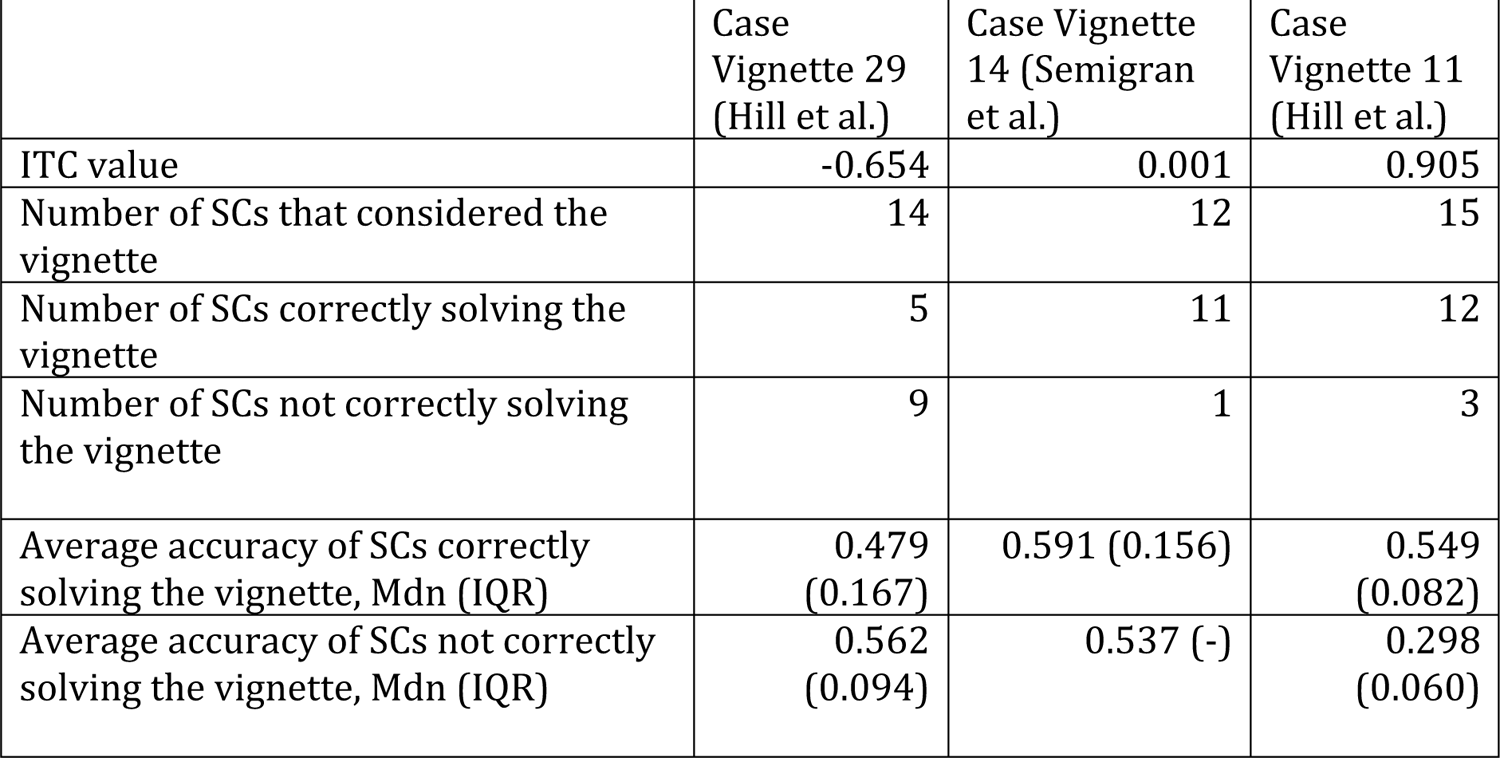
Characteristics of Case Vignettes With The Highest Negative ITC, Lowest ITC (Close to 0) and the Highest Positive ITC

### 3.4. Accuracy Adjusted by Item-Total-Correlation

Adjusting the triage accuracy of symptom checkers by excluding the vignettes with an ITC value below 0.2 changes the accuracy scores of the sample of symptom checkers in both studies: Of the 16 symptom checkers tested by Hill et al., 81% (13/16) showed improvement and 19% (3/16) a decline in accuracy when comparing adjusted to raw accuracy (see Table 4). Their average adjusted accuracy (67.9%, SD = 29.3%) is substantially above the average raw accuracy score reported by Hill et al (48.0%, SD = 10.0%).

**Table 4.** Accuracy of the symptom checkers reported by Hill et al.

| App | Raw Accuracy | Adjusted Accuracy | Rank Based on Raw Accuracy | Rank Based on Adjusted Accuracy |
| --- | --- | --- | --- | --- |
| Healthdirect | 60.9%<br>(28/46) | 90.0%<br>(18/20) | 1 | 4 |
| UofM Health | 57.8%<br>(26/45) | 100.0%<br>(20/20) | 2 | 1 |
| HealthlinkBC | 56.2%<br>(27/48) | 100.0%<br>(21/21) | 3 | 1 |
| Healthwise | 56.2%<br>(27/48) | 100%<br>(21/21) | 3 | 1 |
| Drugs.com<br>Google Play | 56.1%<br>(23/41) | 63.2%<br>(12/19) | 5 | 11 |
| Drugs.com | 53.7%<br>(22/41) | 63.2%<br>(12/19) | 6 | 11 |
| Everyday<br>Health | 51.6%<br>(16/31) | 66.7%<br>(10/15) | 7 | 9 |
| Symptomate | 48.4%<br>(15/31) | 53.3%<br>(8/15) | 8 | 13 |
| Isabel | 47.9%<br>(23/48) | 66.7%<br>(14/21) | 9 | 9 |
| Healthy<br>Children | 47.1%<br>(8/17) | 83.3%<br>(5/6) | 10 | 5 |
| Hopkins all<br>Children | 47.1%<br>(8/17) | 83.3%<br>(5/6) | 10 | 5 |
| CHW | 47.1%<br>(8/17) | 83.3%<br>(5/6) | 10 | 5 |
| St. Lukes<br>Online | 47.1%<br>(8/17) | 83.3%<br>(5/6) | 10 | 5 |
| Symcat | 37.0%<br>(17/46) | 23.8%<br>(5/21) | 14 | 14 |
| Family<br>Doctor | 29.8%<br>(14/47) | 14.3%<br>(3/21) | 15 | 15 |
| Doctor<br>Diagnose | 25.0%<br>(5/20) | 11.1%<br>(1/9) | 16 | 16 |
| Total | 48.0% | 67.8% |  |  |

*Note.* Since the number of entered cases varied for each symptom checker, Total represents the unweighted mean of all symptom checkers.

In contrast, in the Semigran et al study, a minority of 40% (6/15) of audited apps showed an improvement and 60% (9/15) a decline in their accuracy score when comparing adjusted to raw accuracy values (see Table 5). Accordingly, the mean adjusted accuracy (52.8%, SD = 28.4%) was below the mean raw accuracy (58.0%, SD = 12.8%) on a sample level.

**Table 5.** Accuracy of the symptom checkers reported by Semigran et al.

| App | Raw Accuracy | Adjusted Accuracy | Ranked Based on Raw Accuracy | Rank Based on Adjusted Accuracy |
| --- | --- | --- | --- | --- |
| HMS Family Health Guide | 80.0%<br>(32/40) | 95.0%<br>(19/20) | 1 | 2 |
| Healthy Children | 73.3%<br>(11/15) | 100%<br>(8/8) | 2 | 1 |
| Steps2Care | 71.4%<br>(30/32) | 81.0%<br>(17/21) | 3 | 3 |
| Symptify | 70.0%<br>(28/40) | 69.6%<br>(16/23) | 4 | 5 |
| Symptomate | 64.3%<br>(9/14) | 50.0%<br>(2/4) | 5 | 7 |
| Doctor Diagnose | 62.5%<br>(10/16) | 25.0%<br>(1/4) | 6 | 13 |
| Drugs.com | 59.5%<br>(25/42) | 68.2%<br>(15/22) | 7 | 6 |
| FreeMD | 59.1%<br>(26/44) | 72.7%<br>(16/22) | 8 | 4 |
| Family Doctor | 53.7%<br>(22/41) | 50.0%<br>(11/22) | 9 | 7 |
| Early Doc | 52.9%<br>(9/17) | 44.4%<br>(4/9) | 10 | 10 |
| NHS | 52.3%<br>(23/44) | 39.1%<br>(9/23) | 11 | 11 |
| Isabel | 51.1%<br>(23/45) | 30.4%<br>(7/23) | 12 | 12 |
| Symcat | 44.4%<br>(20/45) | 47.8%<br>(11/23) | 13 | 9 |
| Healthwise | 43.2%<br>(19/44) | 18.2%<br>(4/22) | 14 | 14 |
| iTriage | 32.6%<br>(14/43) | 0.0%<br>(0/22) | 15 | 15 |
| Total | 58.0% | 52.8% |  |  |

*Note.* Since the number of entered cases varied for each symptom checker, Total represents the unweighted mean of all symptom checkers.

### 3.5. Item-Total-Correlation by Triage Level

When calculating the item-total-correlation by assessing the correlation of a given vignette with only the subsample of the remaining vignettes of the same triage level, the item-total-correlation values are markedly higher in both studies: Few vignettes in each study show an ITC value of below 0 (Hill: 11.1% (5/45); Semigran: 2.4% (1/41)) or 0.2 (Hill: 20% (9/45); Semigran: 9.7% (4/41)), see Figure 4.

**Figure 4.**
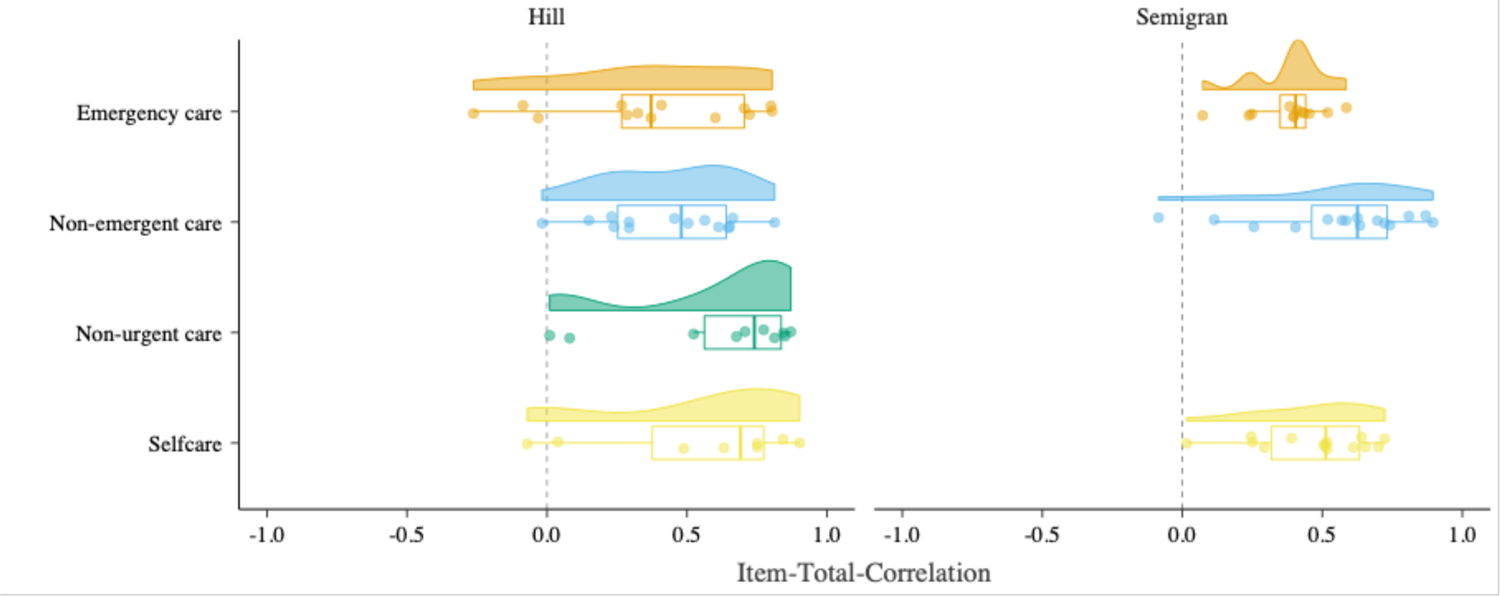
Density of Item-Total-Correlation by Study for Each Triage Level

### 3.6. Comparison of Test Theoretic Metrics of Vignettes Used in Both Studies

The item difficulties of the 30 vignettes used in both studies were moderately correlated (*r* = 0.633, *p* < .001). Item-total-correlations of vignettes used in both studies were correlated negatively, though not reaching statistical significance (r = −0.365, p = 0.073), see Figure 5.

**Figure 5.**
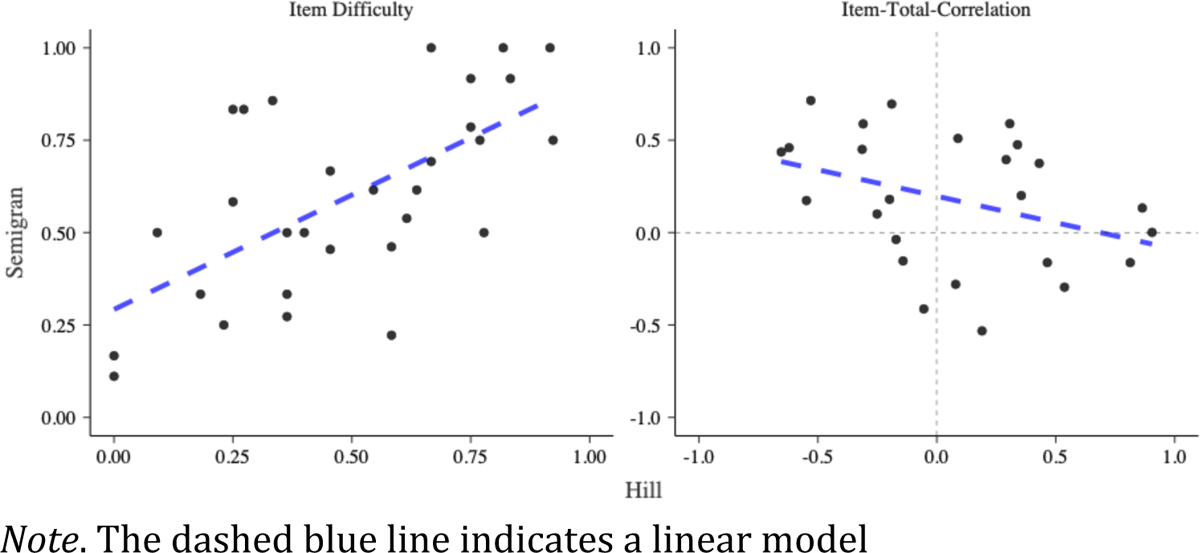
Comparison of Item Difficulty and Item-Total-Correlation of Vignettes Used in Both Studies

## 4. Discussion

### 4.1. Principal Findings

Our findings based on a test theoretic perspective identify previously undescribed limitations of the results reported by two highly influential studies on the capability of symptom checker apps. The calculated metrics, item difficulty (together with the Capability Comparison Score) and item-total-correlation (together with the adjusted accuracy), each provide different but convergent insights.

#### 4.1.1. Item Difficulty

The analysis of item difficulty uncovers that the rank order concerning the competitive comparison of symptom checkers by their raw triage accuracy is easily misleading: Our analyses highlight that apps were assessing not only different numbers and samples of vignettes (that was already reported by Hill et al. and Semigran et al.) but these samples also differed in terms of their difficulty. For example, in the Semigran study the app *Symptomate* was ranked 5 based on (raw) triage accuracy of 64.3% (9/14). The case vignettes it chose to assess, however, were more than 10 percentage points easier (i.e., having a higher item difficulty score) than the mean item difficulty of case vignette samples across all symptom checkers. When considering this by using the CCS to rank order the symptom checkers, *Symptomate*’s rank dropped from 5 to 11 out of 15. With ranks for raw accuracy differing from CSS ranks for a majority of apps in both studies, the validity of the competitive ranking based on accuracy of the symptom checkers is limited. This identified limitation also impacts the interpretation of results on the sample of apps as a whole: The difference between mean raw accuracy and mean CCS values is substantial in the Semigran study. Thus, this study presumably overestimates the capability of the audited symptom checker sample. In contrast, the mean CCS for the sample of apps assessed in the Hill study is similar to their mean raw accuracy score. Consequently, the Hill study’s interpretation of their sample of apps’ triage capability is less influenced by the limitation of apps selectively assessing easier and disregarding harder cases.

#### 4.1.2. Item total correlation

Both assessed studies collected and curated their set of test items (i.e., the case vignettes) purposefully to include a range of rare and common case presentations of different urgency levels, with a gold standard solution provided by a panel of experts. Determining the item-total-correlation of the included case vignette identifies a limitation overlooked by the current method of creating such case vignettes: many vignettes show item total correlation values of below 0.2 making them unsuitable as test items according to common practice in test theory. In practice, one might still consider using vignettes with item total correlation values in the range of 0 to 0.2, if the sample of test items were limited. This would be the case when re-analyzing the performance of symptom checker apps in already published studies using only a few vignettes. For vignette creation in future studies, vignettes with values above 0.2 should be the goal. In our re-analysis, some vignettes even showed a negative item total correlation, that is, overall good apps provided a false recommendation (to this vignette). Thus, based on a test-theoretic perspective, not all vignettes appear to be suitable to evaluate the accuracy of symptom checkers.

Our findings further show reducing the pool of test items (case vignettes) to include only those with an acceptable item total correlation, the results of both studies change: similar to the findings based on item difficulty, both the rank order of individual apps and the performance of the samples of symptom checkers change (with Hill’s study underestimating and Semigran’s study overestimating the capability of their symptom checker sample).

We can only hypothesize as to why such a high proportion of case vignettes achieves only an insufficient item-total-correlation value: First, despite having been reviewed by an expert panel for medical correctness, the respective vignettes may still be ambiguous. That is, they may not include all the necessary information to arrive at the gold standard solution reliably, the gold standard solution may be dependent on context factors such as the healthcare system, which may or may not be considered by the symptom checkers, or the vignettes present information that can be inputted differently in different symptom checkers. For example, the vignette with the lowest ITC (n° 29 in the Hill study) describes a typical presentation of seasonal allergic rhinitis (hay fever) but does not specify whether self-treatment with an over-the-counter antihistamine would be sufficient or non-urgent care should be sought, for example because self-treatment already failed. The negative correlation of item-total-correlation values between the Hill and the Semigran studies supports the hypothesis that ambiguity in the vignettes, inputter variation due to such ambiguity or contextual factors may all compromise the item total correlation of case vignettes.

A further reason for poor ITC values may lie not in the test items (i.e., the case vignettes), but rather in a generalized construct of triage accuracy assumed in most vignette-based studies. That is, many case vignettes achieve only a low ITC value (<0.2) in both studies when considering the entire sample of vignettes. When calculating ITC values for vignettes at the same triage level, however, most vignettes achieved an ITC value greater than 0.2. Thus, case vignettes of a given triage level (e.g., emergency care) may have little or no predictive power on how a symptom checker will perform on case vignettes of a different urgency level (e.g., self-care), but an acceptable predictive power on how the symptom checkers perform on other vignettes of the same triage level. Hence, triage accuracy might be a less valid construct than accuracy per triage level.

A third reason explaining the low ITC values focuses on the symptom checkers, i.e., the “test subjects”. Test theory assumes that test subjects do not err at random. Rather, high-performing test subjects may struggle with difficult test items, but consistently provide correct answers to easy test items. Conversely, low-performing test takers struggle even with easier items and will consistently answer more difficult items incorrectly. However, concerning triage accuracy and symptom checkers, this relationship might be different: a symptom checker might be good at triaging one category of cases (i.e., respiratory tract related cases) but struggle with dermatologic cases, despite most other symptom checkers solving these cases correctly. Figure 1 in the appendix illustrates this: *FamilyDoctor*, one of the poorer performing apps in the Semigran study, is the only app correctly assessing the second most difficult case vignette (the most difficult case vignette was solved correctly by none of the apps). Conversely, *HealthlinkBC* is one of the most capable apps in the Hill study, but the only one incorrectly assessing one of the least difficult case vignettes.

The second and third hypothesized explanations are similar in that they point out that triage capability may not be a unidimensional construct, but in fact it comprises many capabilities. Consequently, rather than assessing symptom checkers’ raw triage accuracy, assessing and comparing them along more differentiated metrics such as sensitivity for emergencies (as proxy for safety) or accuracy per triage level or disease type might provide a more methodologically sound approach to gauge their capability, while the CSS provides the single metric for a more nuanced comparison between symptom checkers.

### 4.2. Lessons a test theory perspective can teach

Case vignette-based studies can only provide limited evidence concerning the accuracy, safety and usefulness of symptom checker apps in comparison to clinical trials ^5^. Nonetheless, they play a vital role in the process of generating evidence: firstly, case vignette-based comparisons of symptom checkers can help to select those best-of-class symptom checkers before testing them in more costly and complex clinical trials. Secondly, symptom checkers as digital tools evolve quickly and their capability and safety may change substantially with each update. Thus, methods with which substantial changes can be noticed without (great) time delay and high costs, like vignette-based assessments, may become important to regulators.

A test theoretic perspective proves useful for both use cases: it can guide the creation of a pool of case vignettes with a balanced spread of item difficulty and high item-total correlation values. Then, few vignettes might suffice as a basis for meaningfully estimating the capability of symptom checkers. This contrasts the current trend of published studies basing their assessment of symptom checkers or differential diagnosis generators on a growing quantity of case vignettes, without scrutinizing their suitability for this purpose ^1, 7, 10, 33, 34^.

Apart from refining the sample of case vignettes (test items), a test theoretic perspective paves the way for both developing more meaningful metrics than raw accuracy values (for example, the CCS presented in this paper), and identifying previously overlooked limitations. We consider these necessary steps towards conducting symptom checker assessments studies “with greater methodological rigour and transparency” as has been recommended ^26^.

### 4.3. Recommendations for future studies on symptom checker accuracy

Based on our results, we recommend that future studies not focus solely on reporting symptom checkers’ accuracy. While this concept remains important and represents an easily comprehensible metric, it is inappropriate to compare the performance of different symptom checkers and seems to be conceptually fuzzy. Instead, reports of accuracy should be restricted to individual symptom checker performance without comparing different symptom checkers and should be accompanied by at least six additional pieces of information: (1) accuracy for different (gold standard) triage levels, (2) safety of advice, (3) comprehensiveness, (4) inclination to overtriage, (5) the item difficulty of different case vignettes used and (6) ranks based on the capability comparison score (CCS). The accuracy for different triage levels allows readers to differentiate between performance in different use cases, e.g., detecting emergencies or advice on whether visiting a professional is indicated at all ^35^. The safety of advice allows readers to assess if symptom checkers give safe advice and can be calculated as the proportion of cases correctly identified as emergencies. The comprehensiveness can give an impression of the breadth of symptoms that can be entered in the symptom checker. It can be calculated as the proportion of case vignettes that could be successfully entered. The inclination to overtriage gives readers an impression on the risk-averseness of the symptom checker and can be calculated as the proportion of cases that received a symptom checker recommendation of higher urgency than the gold standard solution. These metrics allow readers to get an impression of the performance, strengths, and weaknesses of an individual symptom checker. However, these should not be used to compare symptom checkers. Instead, for comparisons, researchers should use item difficulty and the CCS which take into account that different case vignettes were entered and that not all symptom checkers could assess all vignettes. The item difficulty allows readers to infer potential differences in the vignettes that the symptom checkers solved. Lastly, the capability comparison score presented in this paper allows researchers to reliably compare symptom checker performance while accounting for differences in item difficulty and included case vignettes. Thus, this metric enables comparing different symptom checkers without bias due to selective omission of case vignettes. The procedure is visualized in Figure 6.

**Figure 6.**
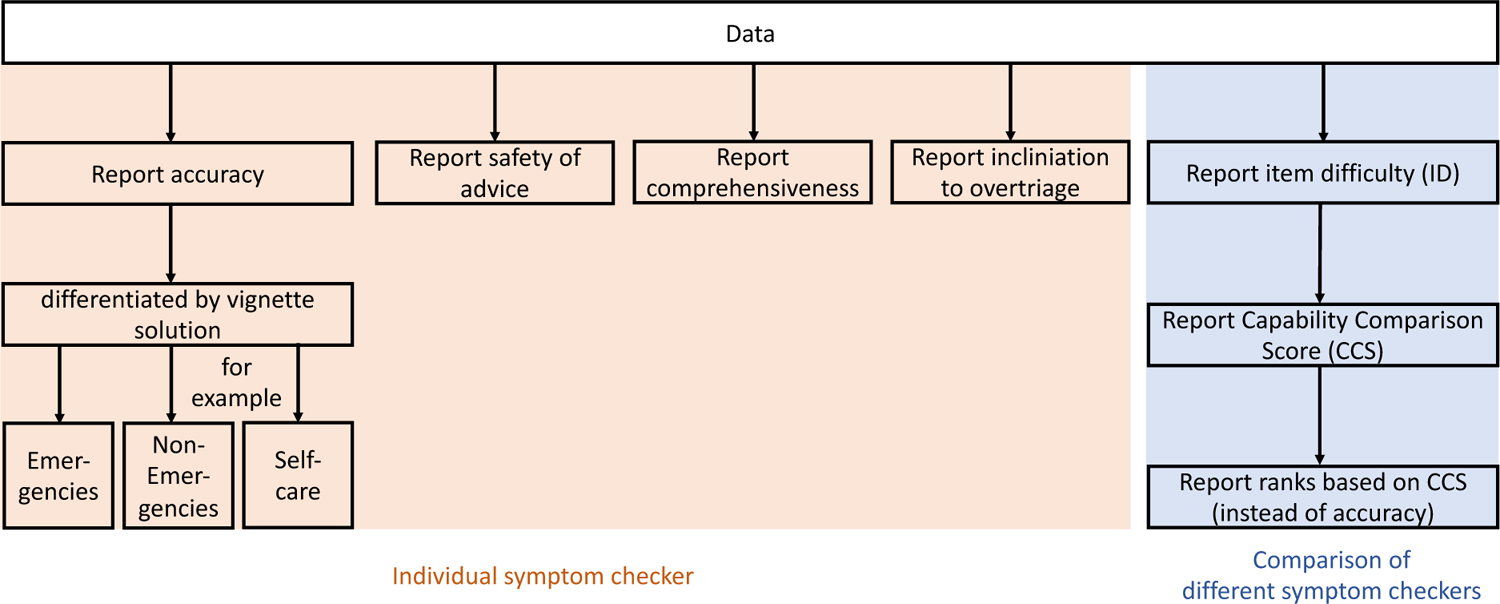
Procedure for Reporting Symptom Checker Performance in Future Studies

Lastly, our findings highlight the importance of sharing data on symptom checker performance among publications: only when authors make data available in such a granular manner that metrics such as item difficulty and ITC can be calculated, can the limitations of their study be truly assessed.

### 4.4. Limitations

We only identified 2 studies that satisfied our inclusion criteria and whose data were available. Thus – although we obtained findings through this initial test theoretic analysis – our results should be validated on other symptom checker evaluation studies. This study also could not include inputter variation, which could explain differences, such as the negative item-total correlation between the two studies. A future study could comparatively enter case vignettes into different symptom checkers and compare item-total correlations between different inputters. Lastly, our study includes metrics from classical test theory to evaluate case vignettes. However, these metrics are not exhaustive and further complementary metrics for the evaluation of case vignette samples - e.g., from item response theory ^36^- are conceivable.

## 5. Conclusions

Applying a test theoretic perspective to two landmark case vignette-based studies, we identified previously unreported limitations in the assessment of symptom checker capabilities. An analysis of item difficulty revealed that many symptom checkers’ triage ability may have been overestimated or underestimated because it did not consider that different symptom checkers may selectively assess easier or more difficult case vignettes. When adjusting for this tendency, the rank order of the symptom checkers differed substantially from the one presented in the original studies. The metric of item total correlation questions the informative value of many case vignettes and/or the metric of (raw) triage accuracy used in these studies.

Although the level of evidence case vignette-based studies can provide is, and will remain, limited, these types of studies will continue to play an important role in the analysis of symptom checkers: to identify symptom checkers that should be tested in more elaborate clinical trials, and as a tool to monitor the development of their growing capability at regular intervals. We propose and define a Capability Comparison Score as a more nuanced metric to compare symptom checkers’ capabilities competitively. The test theoretic perspective presented here is a first step towards improving the theoretical foundations and methodology for benchmarking symptom checkers capabilities with case vignettes, for example by putting more focus on the quality of the case vignettes than their number.

We suggest future studies to present information on the quality and psychometric characteristics of their vignette samples along with their results on the audited symptom checkers.

## Data Availability

Since we did not collect data but used data collected from other authors, we do not publish the dataset used. Generated data can be found in the Appendix.

## Acknowledgements

The authors express their gratitude to Michella G Hill, Hannah L Semigran and their study teams for making their data publicly available.

## Transparency Declaration

The lead author affirms that this manuscript is an honest, accurate, and transparent account of the study being reported, that no important aspects of the study have been omitted and that any discrepancies from the study as planned (and, if relevant, registered) have been explained.

## Abbreviations

ID: Item Difficulty

ITC: Item-Total Correlation

CCS: Capability Comparison Score

## Appendix

**Figure 1 of the Appendix.**
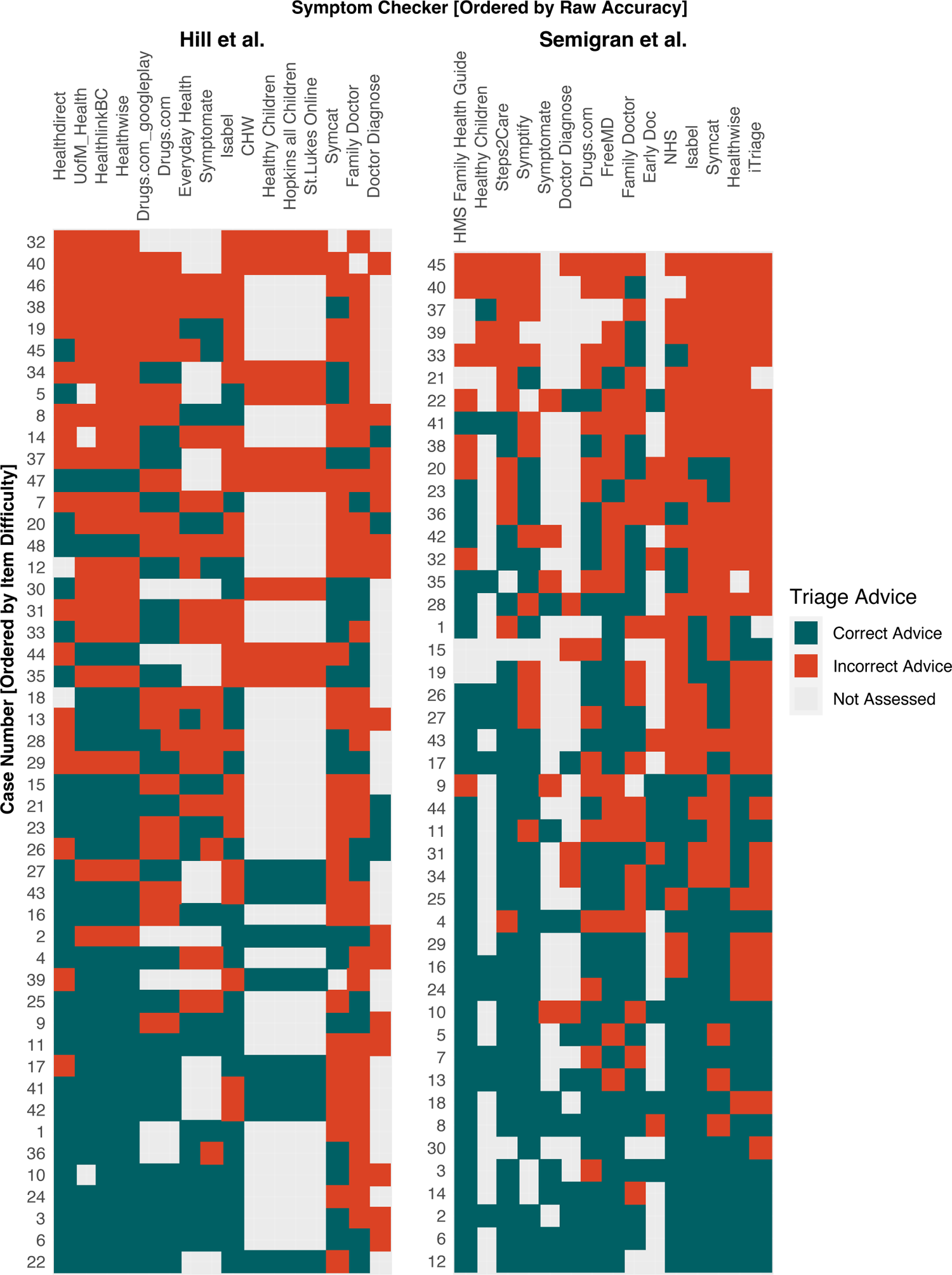

