## Appendix for "How suitable are clinical vignettes for the evaluation of symptom checker apps? A test theoretical perspective"

| **Number** | **Gold Standard Urgency Level** | **Gold Standard Diagnosis** | **Original Text of Case Vignette^3^** | **Semigran et al. Version** | **Semigran et al. Item Difficulty** | **Semigran et al. Item-Total-Correlation** | **Hill et al. Version** | **Hill et al. Item Difficulty** | **Hill et al.**  **Item-Total-Correlation** |
| --- | --- | --- | --- | --- | --- | --- | --- | --- | --- |
| 1 | Em | Acute liver failure (acetaminophen poisoning acceptable) | A 48-year-old woman with a history of migraine headaches presents to the emergency room with altered mental status over the last several hours. She was found by her husband, earlier in the day, to be acutely disoriented and increasingly somnolent. On physical examination, she has scleral icterus, mild right upper quadrant tenderness, and asterixis. Preliminary laboratory studies are notable for a serum ALT of 6498 units/L, total bilirubin of 5.6 mg/dL, and INR of 6.8. Her husband reports that she has consistently been taking pain medications and started taking additional 500 mg acetaminophen pills several days ago for lower back pain. Further history reveals a medication list with multiple acetaminophen-containing preparations. | 48 y/o f, confusion,  disorientation, increasingly  drowsy, mild right upper  quadrant tenderness, chronic  tylenol/acetaminophen -  recently took more | 0.500 | 0.133 | 48-year-old female with history of migraines. Confused. Disoriented. Increasingly sleepy. Mild right upper abdomen tenderness. Has been on various pain medications and has been taking more paracetamol over the last few days because of lower back pain. Chief complaint = confusion | 0.778 | 0.864 |
| 2 | Em | Appendicitis | A 12-year-old girl presents with sudden-onset severe generalised abdominal pain associated with nausea, vomiting, and diarrhoea. On exam she appears ill and has a temperature of 104°F (40°C). Her abdomen is tense with generalised tenderness and guarding. No bowel sounds are present. | 12 y/o f, sudden onset severe  abdominal pain, nausea,  vomiting, diarrhea, T=104 | 1.000 | NA | 12-year-old female. Sudden, severe abdominal pain. Feels sick (nausea). Had vomiting and diarrhoea. Temperature 40°C.  Chief complaint = abdominal pain | 0.667 | -0.294 |
| 3 | Em | Asthma (or acute asthma, status asthmaticus, exacerbation of asthma) | A 27-year-old woman with a history of moderate persistent asthma presents to the emergency room with progressive worsening of shortness of breath, wheezing, and cough over 3 days. She reports prior exposure to a person who had a runny nose and a hacking cough. She did not receive significant relief from her rescue inhaler with worsening symptoms, despite increased use. She has been compliant with her maintenance asthma regimen, which consists of an inhaled corticosteroid and a leukotriene receptor antagonist for maintenance therapy and albuterol as rescue therapy. Her cough is disrupting her sleep pattern and as a consequence she is experiencing daytime somnolence, which is affecting her job performance. | 27 y/o f, Hx of asthma, mild  shortness of breath,  wheezing, 3 days cough,  symptoms not responsive to  inhalers, recent cold | 0.917 | -0.163 | 27-year-old female. History of asthma. Progressively worsening shortness of breath, wheezing and coughing over 3 days. Not responding to inhalers. Cough disturbing her sleep and is sleepy in the daytime. Chief complaint = shortness of breath/breathing problem | 0.833 | 0.813 |
| 4 | Em | COPD exacerbation (severe)  (diagnosis may include acute bronchitis) | A 67-year-old woman with a history of COPD presents with 3 days of worsening dyspnoea and increased frequency of coughing. Her cough is now productive of green, purulent sputum. The patient has a 100-pack-year history of smoking. She has had intermittent, low-grade fever of 100°F (37.7°C) for the past 3 days and her appetite is poor. She has required increased use of rescue bronchodilator therapy in addition to her maintenance medications to control symptoms. | 67 y/o f, Hx of COPD, 3 days  worsening shortness of  breath, increase coughing,  green sputum, low grade  fever, increase use of rescue  bronchodilator therapy | 0.692 | -0.295 | 67-year-old female. History of chronic obstructive airways disease (COPD). 3 days of worsening shortness of breath and cough with green phlegm. Mild fever. (37.7°C) Poor appetite. Increased use of bronchodilator/salbutamol in addition to normal treatment. Smoker (if asked).  Chief complaint = shortness of breath/breathing problem | 0.667 | 0.537 |
| 5 | Em | Deep vein thrombosis | A 65-year-old woman presents with unilateral leg pain and swelling of 5 days' duration. There is a history of    hypertension, mild CHF, and recent hospitalization for pneumonia. She had been recuperating at home but on    beginning to mobilize and walk, the right leg became painful, tender, and swollen. On examination, the right calf is 4    cm greater in circumference than the left when measured 10 cm below the tibial tuberosity. Superficial veins in the    leg are more dilated on the right foot and the right leg is slightly redder than the left. There is some tenderness on    palpation in the popliteal fossa behind the knee. | 65 y/o f, 5 days swelling, pain  in one leg, recent  hospitalization, leg painful,  tender, swollen, red | 0.833 | 0.101 | 65-year-old female. History of high blood pressure and mild congestive heart failure. 5 days of swelling and pain in 1 leg. Recent hospitalisation. Leg is painful, sore, red, swollen.  Chief complaint = Leg pain | 0.273 | -0.251 |
| 6 | Em | Myocardial infarction (heart attack) | Mr. Y is a 64-year-old Chinese male who presents with chest pain for 24 hours. One day before presentation, the patient began to experience 8/10, non-radiating substernal chest pressure associated with diaphoresis and shortness of breath. The pain initially improved with Tylenol, however over the following 24 hours, his symptoms worsened. The patient went to his primary physician, where an EKG was performed which showed ST elevation in leads V2–V6. | 64 y/o m, 1 day chest pain  (8/10), non-radiating  substernal chest pressure,  sweating, shortness of  breath, (chest tightness ) | 1.000 | NA | 64-year-old male. 1 day of chest pain (8/10 pain). Pain does not move elsewhere. Sweating. Breathless. Feels tightness in mid chest (include if asked, as this was an added symptom in Semigran article).  Chief complaint = chest pain | 0.917 | 0.585 |
| 7 | Em | Haemolytic uremic syndrome1 | A 4-year-old boy presents with a 7-day history of abdominal pain and watery diarrhoea that became bloody after the first day. Three days before the onset of symptoms, he had visited the county fair with his family and had eaten a hamburger. Physical examination reveals a mild anaemia. | 4 y/o m, 7 day Hx of  abdominal pain, bloody  diarrhea, ate hamburger at  fair 3 days ago | 0.833 | -0.037 | 4-year-old male. 7 days of stomach pain. Blood in diarrhoea. (Only if asked) - Ate a hamburger at a fair 3 days before stomach-ache began. Chief complaint = diarrhoea | 0.250 | -0.171 |
| 8 | Em | Ureteric stones (kidney stones) | A 45-year-old white man presents to the emergency department with a 1-hour history of sudden onset of left-sided flank pain radiating down toward his groin. The patient is writhing in pain, which is unrelieved by position. He also complains of nausea and vomiting. | 45 y/o m, 1 hour severe leftsided  flank pain radiating into  groin, nausea, vomiting, pain  unrelieved by position | 0.857 | 0.180 | 45-year-old male. 1 hour of severe left sided lower back pain radiating to groin. Feels sick. Vomiting. Pain unrelieved by position. Chief complaint = Back pain | 0.333 | -0.199 |
| 9 | Em | Malaria | A 28-year-old man presents to his physician with a 5-day history of fever, chills, and rigors, not improving with acetaminophen (paracetamol), along with diarrhoea. He had been traveling in Central America for 3 months, returning 8 weeks ago. He had been bitten by mosquitoes on multiple occasions, and although he initially took malaria prophylaxis, he discontinued it due to mild nausea. He does not know the specifics of his prophylactic therapy. On examination he has a temperature of 100.4°F (38°C) and is mildly tachycardic with a BP of 126/82 mmHg. The remainder of the examination is normal. | 28 y/o m, 5 day Hx of fever,  chills, rigors, diarrhea, recent  travel abroad to area with  malaria, bitten by  mosquitoes, did not take  malaria prophylaxis  consistently | 0.583 | -0.413 | 28-year-old male. 5 days of chills, shivering, diarrhoea. (Only if asked by symptom checker - Recently been overseas to New Guinea. Was bitten by mosquitos. Didn’t regularly take anti-malaria medication).  Chief complaint = fever | 0.250 | -0.055 |
| 10 | Em | Meningitis | An 18-year-old male student presents with severe headache and fever that he has had for 3 days. Examination reveals fever, photophobia, and neck stiffness. | 18 y/o m, 3 days severe  headache, fever,  photophobia, neck stiffness | 0.786 | -0.280 | 18-year-old male. 3 days of severe headache. Fever, sore neck and light sensitivity. Chief complaint = headache | 0.750 | 0.079 |
| 11 | Em | Pneumonia | A 65-year-old man with hypertension and degenerative joint disease presents to the emergency department with a  three-day history of a productive cough and fever. He has a temperature of 38.3°C (101°F), a blood pressure of  144/92 mm Hg, a respiratory rate of 22 breaths per minute, a heart rate of 90 beats per minute, and oxygen  saturation of 92 percent while breathing room air. Physical examination reveals only crackles and egophony in the  right lower lung field. The white-cell count is 14,000 per cubic millimeter, and the results of routine chemical tests  are normal. A chest radiograph shows an infiltrate in the right lower lobe. | 65 y/o m, Hx of hypertension  and degenerative joint  disease, 3 day Hx of  productive cough and fever  (101) | 0.615 | -0.162 | 65-year-old male. History of high blood pressure and degenerative joint disease. Had 3 days of coughing up phlegm with a fever. Temperature 38.3°C. Chief complaint = cough | 0.545 | 0.464 |
| 12 | Em | Pulmonary embolism | A 65-year-old man presents to the emergency department with acute onset of short of breath of 30 minutes’ duration. Initially, he felt faint but did not lose consciousness. He is complaining of left-sided chest pain that worsens on deep inspiration. He has no history of cardiopulmonary disease. A week ago, he underwent a total left hip replacement and, following discharge, was on bed rest for 3 days due to poorly controlled pain. He subsequently noticed swelling in his left calf, which is tender on examination. His current vital signs reveal a fever of 100.4°F (38.0°C), heart rate 112 bpm, BP 95/65, and an O2 saturation on room air of 91%. | 65 y/o m, shortness of breath  for 30 min, chest pain that  worsens with inspiration,  recent surgery, recent bed  rest, swelling in left calf,  which is tender, fever | 1.000 | NA | 65-year-old male. Breathless for last 30 minutes. Chest pain on left side which is worse when breathing in. Had recent surgery, with bedrest. Has a swollen left calf which is painful. Has a fever. No history of heart disease.    Chief complaint = shortness of breath/breathing problem | 0.818 | 0.813 |
| 13 | Em | Hendra Virus |  | NA | NA | NA | 29-year-old female. High fever, cough, sore throat, headache and tiredness. Becoming increasingly drowsy. Finding it difficult to breathe. Lives in Queensland and works in stables. | 0.417 | 0.329 |
| 14 | Em | Rocky Mountain Spotted Fever | An 8-year-old boy in Oklahoma is brought to the emergency department over the fourth of July weekend because of  fever, chills, malaise, athralgias, and a headache. Physical examination reveals a maculopapular rash that is most  prominent on his wrists and ankles. | 8 y/o m, Fever, chills, joint  pain, headache, rash  wrists/ankles | 0.846 | 0.134 | NA | NA | NA |
| 15 | Em | Stroke | A 70-year-old man with a history of chronic hypertension and atrial fibrillation is witnessed by a family member to have nausea, vomiting, and right-sided weakness, as well as difficulty speaking and comprehending language. The symptoms started with only mild slurred speech before progressing over several minutes to severe aphasia and right arm paralysis. The patient is taking warfarin. | 70 y/o m, nausea, vomiting,  right-sided weakness, rt arm  paralysis, difficulty speaking  and comprehension | 0.917 | 0.001 | 70-year-old male. History of high blood pressure and atrial fibrillation. Feels sick. Vomiting. Weak down right side. Right arm paralysed. Has trouble speaking and is confused. Takes warfarin. Chief complaint = weak right arm | 0.750 | 0.905 |
| 16 | Em | Tetanus | A 63-year-old man sustained a cut on his hand while gardening. His immunisation history is significant for not having received a complete tetanus immunisation schedule. He presents with signs of generalised tetanus with trismus (“lock jaw”), which results in a grimace described as “risus sardonicus” (sardonic smile). Intermittent tonic contraction of his skeletal muscles causes intensely painful spasms, which last for minutes, during which he retains consciousness. The spasms are triggered by external (noise, light, drafts, physical contact) or internal stimuli, and as a result he is at the risk of sustaining fractures or developing rhabdomyolysis. The tetanic spasms also produce opisthotonos, board-like abdominal wall rigidity, dysphagia, and apnoeic periods due to contraction of the thoracic muscles and/or glottal or pharyngeal muscles. During a generalised spasm the patient arches his back, extends his legs, flexes his arms in abduction, and clenches his fists. Apnoea results during some of the spasms. Autonomic overactivity initially manifests as irritability, restlessness, sweating, and tachycardia. Several days later this may present as hyperpyrexia, cardiac arrhythmias, labile hypertension, or hypotension. | 65 y/o m, cannot open  mouth, contraction of  muscles causing painful  spasms for minutes,  sweating, tachycardia, cut  hand while gardening, did not  get tetanus shot | 0.500 | -0.532 | 65-year-old male. Cannot open mouth. Muscles tightened, causing painful spasms for minutes at a time. Sweating. Fast heart rate. Has cut his hand while gardening and did not get a tetanus injection.  Chief complaint = muscle spasms | 0.364 | 0.190 |
| 17 | NE | Acute otitis media | An 18-month-old toddler presents with 1 week of rhinorrhea, cough, and congestion. Her parents report she is  irritable, sleeping restlessly, and not eating well. Overnight she developed a fever. She attends day care and both  parents smoke. On examination signs are found consistent with a viral respiratory infection including rhinorrhea and  congestion. The toddler appears irritable and apprehensive and has a fever. Otoscopy reveals a bulging,  erythematous tympanic membrane and absent landmarks. | 18 mo f, 1 week rhinorrhea,  cough, congestion, irritable,  lack of appetite, fever, in  daycare | 0.750 | 0.589 | 18-month-old female. 1 week of runny nose, cough, feels chesty, irritable. No appetite. Fever. Goes to day care. Chief complaint = runny nose | 0.769 | 0.307 |
| 18 | NE | Acute pharyngitis | A 7-year-old girl presents with abrupt onset of fever, nausea, vomiting, and sore throat. The child denies cough,  rhinorrhea, or nasal congestion. On physical exam, oral temperature is 101°F (38.5°C) and there is an exudative  pharyngitis, with enlarged cervical lymph nodes. A rapid antigen test is positive for group A Streptococcus (GAS). | 7 y/o f, fever (101), nausea,  vomiting, sore throat, swollen  lymph nodes, tonsilar  exudate; no cough,  rhinorrhea, or nasal  congestion | 0.538 | 0.588 | 7-year-old female. Fever (38.5°C). Feels sick. Vomiting. Sore throat, swollen neck glands. Tonsils have visible pus. No cough, no runny nose nor blocked nose. Chief complaint = fever | 0.615 | -0.310 |
| 19 | NE | Acute pharyngitis | Mr. A is a 24 year-old man who presents to your office for complaints of sore throat, fever, and headache. His  symptoms started 2 days ago with acute onset of sore throat and fever to 102.2. He has had no cough. His physical  examination is normal, except for the presence of tonsillar exudates and some tender anterior cervical  lymphadenopathy. He is otherwise in good health, and is on no medications except for ibuprofen for fever. He has  no drug allergies. (, Centor score = 4 – treat, or test and treat) | 24 y/o m, sore throat, fever  (102.2), headache, no  cough,tonsilar exudates | 0.846 | 0.591 | NA | NA | NA |
| 20 | NE | Acute sinusitis | Mrs. S is a 35 year-old woman who presents with 15 days of nasal congestion. She has had facial pain and green  nasal discharge for the last 12 days. She has had no fever. On physical examination, she has no fever and the only  abnormal finding is maxillary tenderness on palpation. She is otherwise healthy, except for mild obesity. She is on no  medications, except for an over-the-counter decongestant. She has no drug allergies | 35 y/o f, sx for 15 days, nasal  congestion, facial pain, green  nasal discharge, no fever | 0.500 | 0.200 | 35-year-old female. Unwell for 15 days. Has blocked nose with green mucous and facial pain for last 12 days. No fever.  Chief complaint = nasal congestion | 0.400 | 0.355 |
| 21 | NE | Back pain | Consider a 35-year-old man who developed low back pain after shoveling snow 3 weeks ago. He presents to the  office for an evaluation. On examination there is a new left foot drop. In study 82% physicians recommend MRI  (sciatica/sprain) | 35 y/o m, back pain following  shoveling, left foot drop,  symptoms 3 weeks of  duration (loss of sensation in  foot) | 0.333 | -0.153 | 35-year-old male. Back pain following shovelling 3 weeks ago. (Only if asked - Left foot has gone numb and is weak. This was an added symptom in Semigran article). Chief complaint = back pain | 0.182 | -0.142 |
| 22 | NE | Bowel cancer | NA | NA | NA | NA | 56-year-old female with bleeding noticed when opening bowels. Has had some diarrhoea and constipation in the past 6 months. | 0.333 | -0.237 |
| 23 | NE | Cellulitis | A 45-year-old man presents with acute onset of pain and redness of the skin of his lower extremity. Low-grade fever  is present and the pretibial area is erythematous, edematous, and tender. | 45 y/o m, pain and redness of  skin, low grade fever,  redness, edema, and  tenderness lower leg | 0.222 | 0.395 | 45-year-old male. Pain, swollen, sore and redness of skin in lower leg. Mild fever. Chief complaint = leg pain | 0.583 | 0.291 |
| 24 | NE | COPD flare | A 56-year-old woman with a history of smoking presents to her primary care physician with shortness of breath and  cough for several days. Her symptoms began 3 days ago with rhinorrhea. She reports a chronic morning cough  productive of white sputum, which has increased over the past 2 days. She has had similar episodes each winter for  the past 4 years. She has smoked 1 to 2 packs of cigarettes per day for 40 years and continues to smoke. She denies  hemoptysis, chills, or weight loss and has not received any relief from over-the-counter cough preparations. | 56 y/o f, Hx of smoking,  shortness of breath and  cough for several days,  rhinorrhea 3 days ago, white  sputum, no chills | 0.231 | -0.023 | NA | NA | NA |
| 25 | NE | Influenza | A 30-year-old woman presents in January with 2-day history of fever, cough, headache, and generalized weakness.  She was in her usual state of health before an abrupt onset of these symptoms. A few viral illnesses have affected  her during the current winter, but not to this severity. She reports sick contacts at work and did not receive the  seasonal influenza vaccine this season. | 30 y/o f, 2 day fever, cough,  headache, weakness, did not  get flu shot | 0.333 | 0.336 | NA | NA | NA |
| 26 | NE | Mononucleosis | A 16-year-old female high school student presents with complaints of fever, sore throat, and fatigue. She started  feeling sick 1 week ago. Her symptoms are gradually getting worse, and she has difficulty swallowing. She has had a  fever every day, and she could hardly get out of bed this morning. She does not remember being exposed to  anybody with a similar illness recently. On physical examination she is febrile and looks sick. Enlarged cervical lymph  nodes, exudative pharyngitis with soft palate petechiae and faint erythematous macular rash on the trunk and arms  are found. | 16 y/o f, 1 week Hx of fever,  sore throat, fatigue, difficulty  swallowing, fever, enlarged  lymph nodes, exudates,  macular rash on trunk/arms | 0.750 | 0.476 | 16-year-old female. 1-week history of fever, sore throat, fatigue, difficulty swallowing, unable to get out of bed.  Chief complaint = fever | 0.923 | 0.339 |
| 27 | NE | Peptic ulcer disease | A 40-year-old man presents to his primary care physician with a 2-month history of intermittent upper abdominal  pain. He describes the pain as a dull, gnawing ache. The pain sometimes wakes him at night, is relieved by food and  drinking milk, and is helped partially by ranitidine. He had a similar but milder episode about 5 years ago, which was  treated with omeprazole. Physical examination reveals a fit, apparently healthy man in no distress. The only  abnormal finding is mild epigastric tenderness on palpation of the abdomen. | 40 y/o m, 2 month Hx of  intermittent upper abdominal  pain, dulling and gnawing  ache, wakes at night and is  relieved by food/drinking  milk/ranitidine, prior episode  5 yrs ago | 0.667 | 0.509 | 40-year-old male. 2-month history of intermittent upper abdominal pain. (Dull and gnawing ache). Wakes at night and feels better with food, drink, milk or antacid/Gaviscon. Had a similar experience 5 years ago.  Chief complaint = abdominal pain | 0.455 | 0.089 |
| 28 | NE | Pneumonia | A 6-year-old boy with a medical history significant for mild persistent asthma is brought to the clinic by his mother  with a history of a 5-day cough. His mother reports that the child's fever continues to be elevated despite  acetaminophen therapy. He has missed school for the past 3 days and he has a classmate sick with pneumonia. The  mother reports that the appetite is good for the child. His cough produced yellowish sputum at home. His vitals at  the clinic are: respiratory rate 19 breaths/min, heart rate 80 beats/min, and temperature 101.6°F (38.7°C). He  appears in no respiratory distress. His lung examination reveals bilateral rales and occasional wheeze. CXR reveals  lobar infiltrates without pleural effusions. | 6 y/o m, Hx of asthma, 5 days  cough, fever, appetite good,  yellow sputum, t 101.6 | 0.500 | 0.453 | NA | NA | NA |
| 29 | NE | Salmonella | A 14-year-old boy presents with nausea, vomiting, and diarrhea. Eighteen hours earlier, he had been at a picnic  where he ingested undercooked chicken along with a variety of other foods. He reports moderate-volume,  nonbloody stools occurring 6 times a day. He has mild abdominal cramps and a low-grade fever. He is evaluated at  an acute care clinic and found to be mildly tachycardic (heart rate 105 bpm) with a normal BP and a low-grade  temperature of 100.1°F (37.8°C). His physical exam is unremarkable except for mild diffuse abdominal tenderness  and mild increased bowel sounds. He is able to take oral fluids and is instructed on the appropriate oral fluid and  electrolyte rehydration. | 14 y/o m, nausea, vomiting,  non-bloody diarrhea, mild  abdominal cramps (T=100.1),  mild abdominal tenderness,  diarrhea after attending a  picnic and eating  undercooked chicken, | 0.500 | 0.374 | NA | NA | NA |
| 30 | NE | Migraine | NA | NA | NA | NA | 44-year-old female. 2-day severe throbbing headache. Feels nauseated when she moves.  Wants to stay in bed with curtains closed. Family has history of migraines. | 0.583 | 0.291 |
| 31 | NE | Queensland tick typhus | NA | NA | NA | NA | 25-year-old female. Been unwell for 9 days. Fever (38.5°C), headache, dry cough, widespread rash. Muscle weakness. Painful upper left abdomen, with nausea and vomiting. Painful joints. Has been camping and gone on bush walks. | 0.818 | 0.869 |
| 32 | NE | Ross River virus | NA | NA | NA | NA | 19-year-old male. 2 weeks of fever with chills, muscle aches and joint pain with swelling and  stiffness at joints. Rash. Fatigue. Swollen glands. Headache behind the eyes. | 0.727 | 0.210 |
| 33 | NE | Shingles | A 77-year-old man reports a 5-day history of burning and aching pain on the right side of his chest. This is followed  by the development of erythema and a maculopapular rash in this painful area, accompanied by headache and  malaise. The rash progressed to develop clusters of clear vesicles for 3 to 5 days, evolving through stages of  pustulation, ulceration, and crusting. | 77 y/o m, 5 day burning and  aching on right side of chest,  erythema, maculopapular  rash, headache, malaise, rash  progressed to clear vesicles  after 3-5 days | 0.462 | 0.450 | 7-year-old male. 5 days of burning and pain on right side of chest. Chest is red with a rash, some spots are clear raised bumps, while some are red and flat. Has a headache and feels tired and unwell.  Chief complaint = chest pain | 0.583 | -0.314 |
| 34 | NE | Urinary tract infection | A 26-year-old female newly wed presents complaining of painful urination, feeling of urgent need to urinate, and  more frequent urination for 2 days. She denies any fever, chills, nausea, vomiting, back pain, vaginal discharge, or  vaginal pruritus. | 26 y/o f, painful urination,  urgent need to urinate, more  frequent urination for 2 days,  sexually active; no fever,  chills, nausea, vomiting, back  pain, vaginal discharge,  vaginal pruritus | 0.727 | 0.572 | NA | NA | NA |
| 35 | NE | Vertigo | A 65-year-old woman presents with a chief complaint of dizziness. She describes it as a sudden and severe spinning  sensation precipitated by rolling over in bed onto her right side. Symptoms typically last <30 seconds. They have  occurred nightly over the last month and occasionally during the day when she tilts her head back to look upward.  She describes no precipitating event prior to onset and no associated hearing loss, tinnitus, or other neurologic  symptoms. Otologic and neurologic examinations are normal except for the Dix-Hallpike maneuver, which is negative  on the left but strongly positive on the right side. | 5 y/o f, dizziness, sudden  onset, recurrent, lasts <30  sec, consistent trigger, no  hearing loss, ringing in ears,  muscle weakness, loss of  sensation | 0.889 | 0.539 | NA | NA | NA |
| 36 | NE | Bursitis (trochanteric bursitis) | NA | NA | NA | NA | 60-year-old woman. Developed pain in L hip, hurts to sleep on that side. Otherwise well. | 0.364 | 0.231 |
| 37 | NE | Molluscum contagiosum | NA | NA | NA | NA | 7-year-old male. Has small raised shiny pearly spots on his stomach and back with a dot in the  middle. They are not painful. (If asked, no fever  or other illness. Not itchy.) | 0.385 | -0.365 |
| 38 | NE | Solar keratosis | NA | NA | NA | NA | 43-year-old female. She has fair skin. Has a small scab (3 mm) on top of head which grows back if knocked off. Dry and rough to touch. | 0.778 | 0.495 |
| 39 | NE | Sprained ankle | NA | NA | NA | NA | 10-year-old female. Fell and twisted ankle. Slightly swollen and bruised ankle. Limping, and ankle is sore. No deformity, no sound of broken bones. | 0.286 | -0.352 |
| 40 | S-c | Acute bronchitis | A 34-year-old woman with no known underlying lung disease 12-day history of cough. She initially had nasal  congestion and a mild sore throat, but now her symptoms are all related to a productive cough without paroxysms.  She denies any sick contacts. On physical examination she is not in respiratory distress and is afebrile with normal  vital signs. No signs of URI are noted. Scattered wheezes are present diffusely on lung auscultation. | 34 y/o f, 12 day cough, initial  nasal congestion and sore  throat, cough, no fever | 0.615 | 0.375 | 34-year-old female. 12 days of coughing. Initially had blocked nose and sore throat. Now has cough which brings up phlegm, but no fever.  Chief complaint = cough | 0.636 | 0.431 |
| 41 | S-c | Acute bronchitis | Mrs. L is a 61 year-old woman who presents with 4 days of a cough productive of yellow sputum. Her symptoms  started 4 days ago with rhinorrhea and productive cough. She initially had fevers as high as 101 for 2 days, but those  have now resolved. In the office, she has normal vital signs and a normal physical examination. She is otherwise  healthy except for high cholesterol for which she is being treated with atorvastatin. She has no drug allergies. | 61 y/o f, 4 day cough, yellow  sputum, rhinorrhea, fever  (resolved) | 0.417 | 0.274 | NA | NA | NA |
| 42 | S-c | Acute conjunctivitis | A 14-year-old boy with no significant past medical history presents 3 days after developing a red, irritated right eye  that spread to the left eye today. He has watery discharge from both eyes and they are stuck shut in the morning. He  reports recent upper respiratory symptoms and that several children at his day camp recently had pink eye. He  denies significant pain or light sensitivity and does not wear contact lenses. On examination, his pupils are equal and  reactive and he has a right-sided, tender preauricular lymph node. Penlight examination does not reveal any corneal  opacity. | 14 y/o m, 3 days red, irritated  eye (spread from right to  left), discharge, URI  symptoms, no pain or light  sensitivity | 0.167 | -0.217 | 14-year-old male. 3 days with red irritated eye (spread from right to left eye). Eyes have pus from ducts. Has recently had cold like symptoms. No pain or light sensitivity.  Chief complaint = red eye | 0.000 | NA |
| 43 | S-c | Cradle cap | NA | NA | NA | NA | 4-week-old male. Crusty yellowish skin appearing on head and behind ears, which looks waxy. | 0.769 | 0.611 |
| 44 | S-c | Dysmenorrhoea | NA | NA | NA | NA | 15-year-old female. Monthly mid-abdominal pain appears with period. Pain radiates to back. Headache. | 0.769 | 0.611 |
| 45 | S-c | Head lice | NA | NA | NA | NA | 4-year-old female. Has very itchy head. Can see small white dots in her hair near scalp, especially behind ears. Some white dots are on hair strands and don’t brush off. | 0.615 | 0.249 |
| 46 | S-c | Herpes simplex virus type 1 (cold sore) | NA | NA | NA | NA | 17-year-old male. Blisters appearing on lips and just inside of mouth. Before blisters appeared, he felt tingling and was itchy. | 0.364 | -0.050 |
| 47 | S-c | Plantar warts | NA | NA | NA | NA | 20-year-old male. Hard skin-coloured lumps on the sole of the foot. Painful to walk on. Some have a black dot in the centre. | 0.182 | 0.108 |
| 48 | S-c | Threadworm (Strongyloides) | NA | NA | NA | NA | 5-year-old male. Itchy bottom. White threads appearing in stools. Grumpy and tired. | 0.286 | 0.517 |
| 49 | S-c | Tinea pedis (athlete’s foot) | NA | NA | NA | NA | 33-year-old male. Scaly skin between the toes. The skin is Itchy and has an odour. Skin goes soft and white when wet. | 0.333 | 0.526 |
| 50 | S-c | Acute pharyngitis | Mr. E is a 26 year-old man who presents to your office for complaints of sore throat, headache, and non-productive  cough. His symptoms started 2 days ago with acute onset of sore throat. He has been afebrile. His physical  examination is normal, except for some pharyngeal erythema. He is otherwise in good health, and is on no  medications except for acetaminophen for his sore throat and fever. He has no drug allergies. | 26 y/o m, 2 day sore throat,  headache, cough, no fever | 0.615 | 0.370 | NA | NA | NA |
| 51 | S-c | Acute rhinitis | A 22-year-old student presents with a 5-year history of worsening nasal congestion, sneezing, and nasal itching.  Symptoms are year-round but worse during the spring season. On further questioning it is revealed that he has  significant eye itching, redness, and tearing as well as palate and throat itching during the spring season. He  remembers that his mother told him at some point that he used to have eczema in infancy. | 22 y/o m, 5 year Hx of nasal  congestion, sneezing, nasal  itching worse during spring  season, eye itching, redness,  tearing, palate and throat  itching, Hx of eczema in  infancy | 0.455 | 0.436 | 22-year-old male. 5-year history of blocked nose, sneezing, nasal itching which is worse in spring. Has itchy eyes which are red and watery. Throat and palate are also itchy. Has a history of eczema in early childhood. Chief complaint = blocked nose | 0.455 | -0.654 |
| 52 | S-c | Back pain | A 38-year-old man with no significant history of back pain developed acute LBP when lifting boxes 2 weeks ago. The  pain is aching in nature, located in the left lumbar area, and associated with spasms. He describes previous similar  episodes several years ago, which resolved without seeing a doctor. He denies any leg pain or weakness. He also  denies fevers, chills, weight loss, and recent infections. Over-the-counter ibuprofen has helped somewhat, but he  has taken it only twice a day for the past 3 days because he does not want to become dependent on painkillers. On  examination, there is decreased lumbar flexion and extension secondary to pain, but a neurologic exam is  unremarkable. | 38 y/o m, acute low back pain  after lifting, no leg pain or  weakness, no fevers, chills,  weight loss, or recent  infections | 0.333 | 0.459 | 38-year-old male. Sudden lower back pain after lifting. No leg pain or weakness, no fevers or chills, no weight loss or recent infections. Chief complaint = back pain | 0.364 | -0.621 |
| 53 | S-c | Bee sting | A 9-year-old boy is brought to the ER after being stung by a bee at a picnic. He is crying hysterically. After 15 minutes  of calming him down, exam reveals a swollen tender upper lip but no tongue swelling, no drooling, no stridor, no  rash, and no other complaints. | 9 y/o m, bee sting, swollen  and tender upper lip; no  tongue swelling, drooling,  stridor, rash, or other  complaints | 0.111 | 0.425 | 9-year-old male. Stung by a bee, swollen and sore upper lip. No tongue swelling, drooling, noisy breathing, rash or other complaints.  Chief complaint = bee sting | 0.000 | NA |
| 54 | S-c | Canker sore | A 17-year-old male student presents with recurrent mouth ulceration since his early schooldays. He has no  respiratory, anogenital, gastrointestinal, eye, or skin lesions. His mother had a similar history as a teenager. The  social history includes no tobacco use and virtually no alcohol consumption. He has no history of recent drug or  medication ingestion. Extraoral exam reveals no significant abnormalities and specifically no pyrexia; no cervical  lymph node enlargement; nor cranial nerve, salivary, or temporomandibular joint abnormalities. Oral exam reveals a  well-restored dentition and there is no clinical evidence of periodontal-attachment loss or pocketing. He has five 4  mm round ulcers with inflammatory haloes in his buccal mucosae. | 17 y/o m with recurrent  mouth ulceration for year, no  respiratory, anogenital,  gastrointestinal, eye, or skin  lesions, mother has similar  Hx, no Hx of recent drugs or  medication | 0.273 | 0.174 | 17-year-old male. Reoccurring mouth ulcers for a year. No respiratory, anal or genital, gastrointestinal, eye or skin lesions. Mother has similar history. No history of drugs or medications.  Chief complaint = mouth ulcers | 0.364 | -0.548 |
| 55 | S-c | Candidal yeast infection | Consider a 40-year-old, monogamous, married woman who calls to report a 2-day history of vaginal itching and  thick white discharge. She has no abdominal pain or fever. (in study 50% recommended physician visit) | 40 y/o f, 2 day vaginal itching,  thick white discharge, no  abdominal pain or fever | 0.111 | -0.062 | NA | NA | NA |
| 56 | S-c | Constipation | A 5-month-old baby boy presents with difficulty and delay in passing hard stools. His mother reports that he strains  for several hours and may even miss a day, before passing stool with screaming and occasional spots of fresh blood  on the stool or diaper. He has recently been weaned from breastfeeding to cows' milk formula, which he had been  reluctant to drink initially. The child is thriving and now feeding normally. There was no neonatal delay in defecation  and no history of excessive vomiting or abdominal distension. | 5 mo m, difficulty/delay in  passing hard stools, strains  for hours, may miss a day,  screams when passes stool  and occasional spots of  blood, weaned from  breastmilk to cows' milk, now  feeding normally | 0.091 | -0.149 | NA | NA | NA |
| 57 | S-c | Eczema | A 12-year-old female presents with dry, itchy skin that involves the flexures in front of her elbows, behind her knees,  and in front of her ankles. Her cheeks also have patches of dry, scaly skin. She has symptoms of hay fever and has  recently been diagnosed with egg and milk allergy. She has a brother with asthma and an uncle and several cousins  who have been diagnosed with eczema. | 12 y/o f, dry, itchy skin in  front of elbows, behind  knees, in front of ankles,  cheeks have patches of dry,  scaly skin, symptoms of hay  fever, egg and milk allergy,  brother has asthma and uncle  and cousins have eczema | 0.250 | 0.696 | 12-year-old female. Dry, itchy skin in front of elbows, in front of knees and cheeks have patches of dry, scaly skin. Symptoms of hay fever. Has egg and milk allergy. Brother has asthma and Uncle and cousins have eczema.  Chief complaint = rash | 0.231 | -0.190 |
| 58 | S-c | Stye | A 30-year-old man presents with a painful, swollen right eye for the past day. He reports minor pain on palpation of  the eyelid and denies any history of trauma, crusting, or change in vision. He has no history of allergies or any eye  conditions and denies the use of any new soaps, lotions, or creams. On exam, he has localized tenderness to  palpation and erythema on the midline of the lower eyelid near the lid margin. The remainder of the physical exam,  including the globe, is normal. | 30 y/o m, painful, swollen  right eye for past day, no Hx  of trauma, crusting, change in  vision, allergies, or eye  conditions, localized  tenderness, erythema  (redness) | 0.333 | 0.423 | NA | NA | NA |
| 59 | S-c | Viral upper respiratory tract infection | Mr. R. 5is a 56 year-old man who presents to you with 6 days of non-productive cough, nasal congestion, and green    nasal discharge. He has had intermittent fevers as high as 100.8. His physical examination is normal except for    rhinorrhea. He is otherwise healthy, except for chronic osteoarthritis of the right knee. He has no drug allergies. | 56 y/o m, 6 day cough, nasal  congestion, green nasal  discharge, fever (100.8),  rhinorrhea | 0.500 | 0.715 | 56-year-old male, 6-day cough, nasal congestion, green nasal discharge. Fever (38.2°C) and runny nose.  Chief complaint = cough | 0.091 | -0.530 |
| 60 | S-c | Viral upper respiratory tract infection | A 30-year-old man presents with a 2-day history of runny nose and sore throat. He feels hot and sweaty, has a mild  headache, is coughing up clear sputum and complains of muscle aches. He would like antibiotics as he was  prescribed them last year when he had a similar condition. On examination, he is afebrile, has a normal pulse, a  slightly inflamed pharynx and nontender cervical lymphadenopathy. There is no neck stiffness and his chest is clear.  He has tried over-the-counter cough medications, but has not found these helpful. He smokes 10 cigarettes per day. | 30 y/o m, 2 day HX of runny  nose, sore throat, hot,  sweaty, mild headache, cough  with clear sputum, muscle  aches, no fever or neck  stiffness | 0.583 | 0.428 | NA | NA | NA |
| 61 | S-c | Vomiting | Elizabeth’s 2-year-old son has a fever and vomited twice. Elizabeth worries about dehydration, so she gives Jack a  sippy cup of apple juice. He immediately vomits up the juice. Elizabeth debates what to do next. Should she try to  reach Jack’s pediatrician or should she take Jack to the ED? Instead, she calls her triage nurse line. Temperature =  100.5 | 2 y/o m, low grade fever (T =  100.5), vomited twice, vomits  up juice | 0.000 | NA | NA | NA | NA |
